# Antibiotic Utilisation Patterns in Tanzania: A Retrospective Longitudinal Study Comparing Pre- and Post-COVID-19 Pandemic Using Tanzania Medicines and Medical Devices Authority Data

**DOI:** 10.1101/2023.11.27.23299060

**Authors:** Raphael Z. Sangeda, Sahani M. William, Faustine Cassian Masatu, Adonis Bitegeko, Yonah Hebron Mwalwisi, Emmanuel Alphonse Nkiligi, Pius Gerald Horumpende, Adam M. Fimbo

## Abstract

**Background:** Antimicrobial resistance (AMR) is a growing public health concern globally, and misuse of antibiotics is a major contributor.

**Objective:** This study investigated antibiotic utilisation patterns before and after the COVID-19 pandemic in Tanzania using data from the Tanzania Medicines and Medical Devices Authority (TMDA).

**Methods:** This retrospective longitudinal study analysed secondary data. The study compared antibiotics consumption in defined daily doses (DDD) per 1000 inhabitants per day (DID) in two distinct eras: 2018-2019 as the pre-COVID-19 era and 2020-2021 as the post-COVID-19 era. Data was reorganised using Microsoft Power BI, and statistical analysis was conducted using SPSS software.

**Results:** The study analysed 10,614 records and found an overall increase in antibiotics consumption from 2018 to 2021. When we divided the consumption of antibiotics into a pre- and post-COVID time period, with the pre-COVID period being 2018 and 2019 and the post-COVID period being 2020 and 2021, we found that the consumption was 61.24 DID in the post-COVID era and 50.32 DID in the pre-COVID era. Levofloxacin had the highest percentage increase in use, with a 700% increase in DID after the pandemic. Azithromycin had a 163.79% increase, while cefotaxime had a 600% increase. In contrast, some antibiotics exhibited a decrease in usage after the pandemic, such as nalidixic acid, which had a 100% decrease, and cefpodoxime, 66.67% decrease.

**Conclusion:** The increase in antibiotic consumption during the COVID-19 pandemic highlights the importance of implementing effective antimicrobial stewardship strategies to prevent AMR, especially during pandemics.

## Introduction

Antimicrobial resistance (AMR) poses a serious global health threat by hindering the treatment of bacterial infections.^1^ In low and middle-income countries (LMICs), Tanzania included, the misuse and overuse of antibiotics have resulted in high rates of AMR, making it challenging to treat bacterial infections^2,3^. Proper regulation and minimal consumption of antibiotics are crucial in reducing the prevalence of AMR in LMICs^2^.

To increase healthcare professionals’ capacity for health safety and security and provide awareness on the appropriate use of antibiotics to reduce the development of AMR, global partnerships such as the Global Health Security Agenda (GHSA) and Antimicrobial Stewardship Programs (ASP) have been created. However, the COVID-19 pandemic has led to increased use of antibiotics due to the absence of antiviral therapies, which may contribute to further development of AMR. The second reason for increased antibiotics use has been the lack of proper infection prevention control (IPC) measures in public areas and hospitals.^4^

The government of Tanzania has implemented various programs to provide affordable healthcare services to the population, including a pre-payment system and health insurance scheme. Further, the Parliament of Tanzania has recently passed a bill for universal health insurance, awaiting the President’s signature into law. However, achieving universal health coverage in Tanzania remains challenging as many citizens still lack access to proper healthcare services and have access to antibiotics without a prescription.^5^

The World Health Organization (WHO) has promulgated several guidelines for countries embarking on AMS to optimise antibiotics prescription and dispensing. One such guideline is using the AWaRe (Access, Watch, Reserve) classification of antibiotics in reporting AMU data.^6^ According to this AWaRe classification, the Access group consists of antibiotics with activity against a wide range of commonly encountered susceptible bacteria with lower resistance potential than antibiotics in the other groups. The Watch class consists of antibiotics with higher resistance potential and includes most of the highest priority agents among the critically important antimicrobials for human use. The Reserve group includes antibiotics and antibiotic classes that should be reserved to treat confirmed or suspected infections due to multi-drug-resistant organisms. Reserve group antibiotics should be treated as “last resort” options.^7,8^ This classification emphasises using the reserve group sparingly to preserve strong antibiotics for serious infections. Other guidelines include the Anatomical Therapeutic Chemical (ATC) classification system and the daily defined dose (DDD) system to allow for international standardisation in AMU studies and permit comparison across geographical regions.^9^

At the onset of the COVID-19 outbreak in Tanzania in March 2020^10,11^ clinicians began administering antibiotics to patients presenting with clinical signs such as cough, fever, and radiological infiltrate indicative of bacterial community-acquired pneumonia. The absence of antiviral therapies with proven efficacy also contributed to the widespread and excessive prescription of antibiotics, leading to increased antimicrobial resistance during the pandemic^11–13^.

Implementing proper antibiotic stewardship programs and increasing awareness of the appropriate use of antibiotics among healthcare professionals and the general public is pivotal in combating AMR. Collaboration among stakeholders is necessary to ensure effective treatment of bacterial infections to combat AMR.^14^

In Tanzania, AMS efforts underpinned in the National Action Plan (NAP) were launched^15,16^ in 2017 and the second version^17^ in 2022 to help combat AMR. In 2020, the emergence of the COVID-19 pandemic impacted many aspects of life, including antimicrobial use (AMU) in Tanzania and globally. However, there is a paucity of AMU data in Tanzania comparing pre and post-COVID-19 eras to guide AMS. The Tanzania Medicines and Medical Devices Authority (TMDA) regulates the use of antibiotics in the country and collects data on their utilization^15^. TMDA’s data on antibiotics utilisation can provide valuable information to develop effective strategies to combat AMR in Tanzania. Therefore, this study aimed to investigate the change in antibiotic utilisation patterns before and after the COVID-19 pandemic in Tanzania, using TMDA data from 2018 to 2021 to determine the trends of use before and during the COVID-19 pandemic outbreak in Tanzania.

## Methods

### Study design

This retrospective and longitudinal study used data from the Tanzania Medicines and Medical Devices Authority (TMDA), covering four years from 2018 to 2021.

### Study area

The study was conducted in Tanzania Mainland, and the data were collected from TMDA headquarters in Dodoma. The TMDA compiled all importation data at different ports of entry from January 2018 to December 2021, which were included in this study.

### Inclusion criteria

The study included all records of antibiotics for intended human use administered systemically.

### Exclusion criteria

Antifungals, antivirals, antiparasitic and topical antibiotics and non-antibiotics were excluded from the study.

### Data collection

Data were obtained from the TMDA headquarters in Dodoma, where individual importation data were recorded from January 2018 to December 2021. The data collected include product descriptions, generic and trade names, strengths, dosage forms, pack sizes, price, quantities, unit prices, import permits identification, issue dates, supplier names, countries, and local importers.

The TMDA is the National Medicines Regulatory Authority (NMRA) responsible for regulating the importation of medicines into the Tanzanian mainland market. The TMDA has developed and issued regulations and procedures that compel importers to apply for an importation permit. Importers’ applications are evaluated, and import permits are issued and archived in the Regulatory Information Management System (RIMS), from which the records of imported medicines can be retrieved ^15^. The data included antibiotic generic name, country of origin, ATC class, imports, and WHO AWaRe classification status of antibiotics received. The WHO ATC classification system categorised antibiotics into their respective classes.

Results were expressed using the Daily Defined Doses (DDD) measurement units. Utilisation was expressed in DDD per 1000 inhabitants per day (DID) in accordance with the ATC/DDD WHO collaborating Center for Statistics Methodology ^18^.

### Data analysis

Data collected were checked for completeness, accuracy, and omissions using Microsoft Power BI. Homogeneous data were clustered and coded for easy analysis, and the findings were presented in tables and graphs. Statistical analysis such as paired samples t-test was conducted in Statistical Package for the Social Sciences (SPSS) version 26.0 to assess the impact of the pre-and post-COVID-19 pandemic on antibiotics consumption in Tanzania. Time series and regression analyses were performed to predict the annual trend of antibiotic utilisation. An autoregressive integrated moving average (ARIMA, 0, 1, 0) model was established to predict the trends of antibiotic use. A *p*-value of less than 0.05 was considered statistically significant.

### Ethical considerations

Ethical clearance was obtained from the Directorate of Research and Publications of the Muhimbili University of Health and Allied Sciences (MUHAS) with reference number DA. 25/111/28/01/2021. The TMDA granted permission to collect and use its accrued data. Data privacy and confidentiality were observed throughout the study. Patient identifying information was not collected since this did not involve direct patient contact; instead, secondary data from the TMDA importation records was used.

## Results

A total of 10,627 records were retrieved from the TMDA information management system. Of these, 9,610 were antibiotics imported for systemic use in humans between 2018 and 2021. A total of 1,017 records were excluded because they referred to antibiotics for either topical or veterinary use, as they were non-J (01) level 2 ATC class and 2022 imports. A total of 117.02 DID was utilised in Tanzania between 2018 and 2021 (Table 1), with a mean (standard deviation) of 29.25 (±4.63) DIDs.

**Table 1:** Annual distribution of DIDs and number of permits of antibiotics imported in Tanzania between 2018 and 2021.

| Year | DID | Number of Permits |
| --- | --- | --- |
| 2018 | 30.39831 | 2,491 |
| 2019 | 22.53625 | 2,426 |
| 2020 | 30.96806 | 2,152 |
| 2021 | 33.11989 | 2,541 |
| **Total** | **117.0225** | **9,610** |

Key: DID: Daily Defined Dose per 1000 inhabitants per day

The year 2021 had the highest DID at 33.1, 47.0% higher than 2019, with the lowest DID at 22.5. The consumption in 2021 accounted for 28.30% of DIDs. Annual cumulative DID started shooting up in 2020, where cumulative yearly DID reached 83.9 of the total 117.02 for the four years.

Tanzania imports these antibiotics from across continents and Kenya, India, and China were the major sources of antibiotics in the pre- and post-COVID-19 eras. Tanzania and South Africa were sources of antibiotics only during the pandemic era (Figure 1).

**Figure 1:**
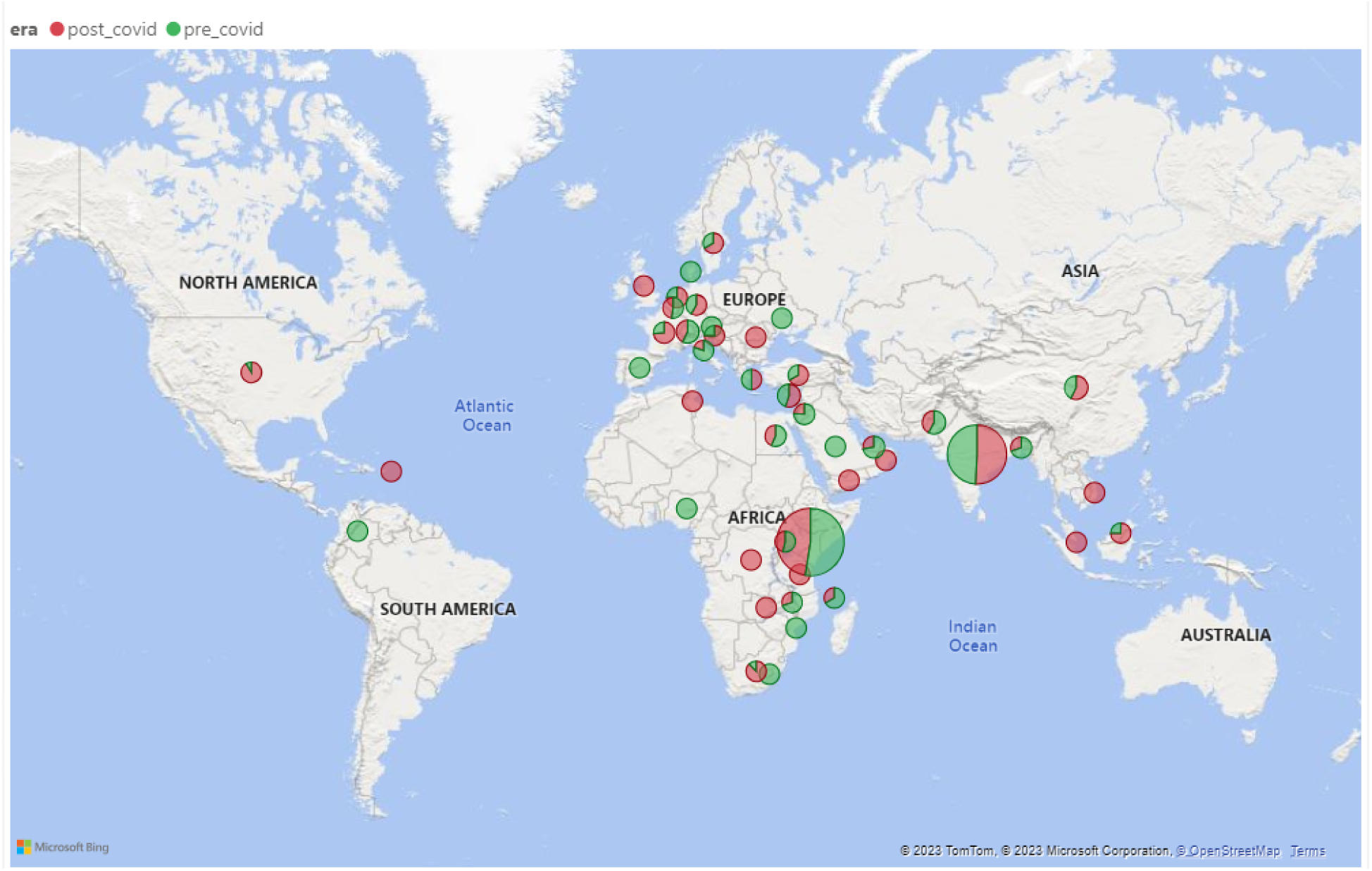
Worldwide country frequency of importations pre-and post-COVID-19 pandemic. The size of the bubble signifies the frequency contributed by the respective country.

The oral and parenteral dosage forms contributed 151.18 (96.93%) and 3.33 (3.075%) of the DIDs, respectively (Figure 2). The contribution of individual dosage forms indicates that capsules contributed the most (Supplementary Figure 1) and Supplementary Table 1.

**Figure 2:**
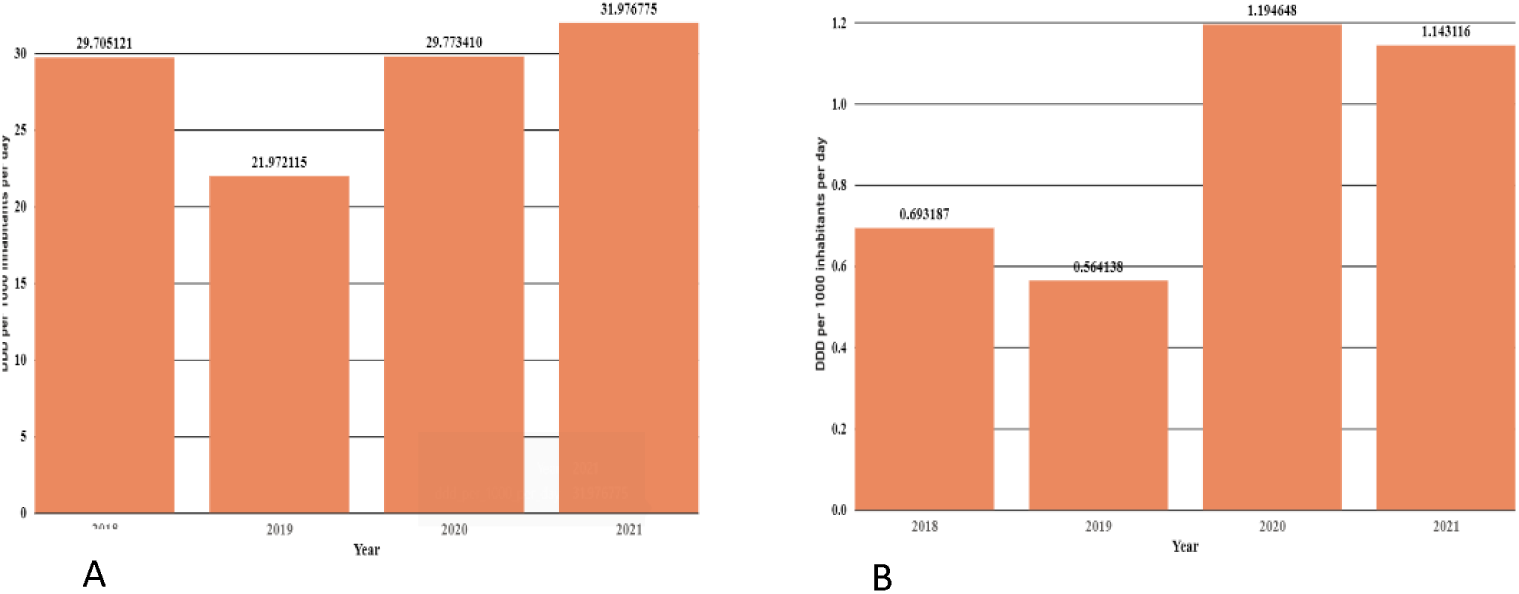
DID contribution for oral (Panel A) and parenteral (panel B) antibiotics.

Overall, the Access group had the highest DDI at 82.9, followed by Watch, other, and Reserve. Access group accounted for 70.8% (Figure 3) of DDI, while the WHO suggests > 60%. The increase in the Watch group of antibiotics parallels a general decline in the Access group (Figure 3) and (Supplementary Figure 2).

**Figure 3:**
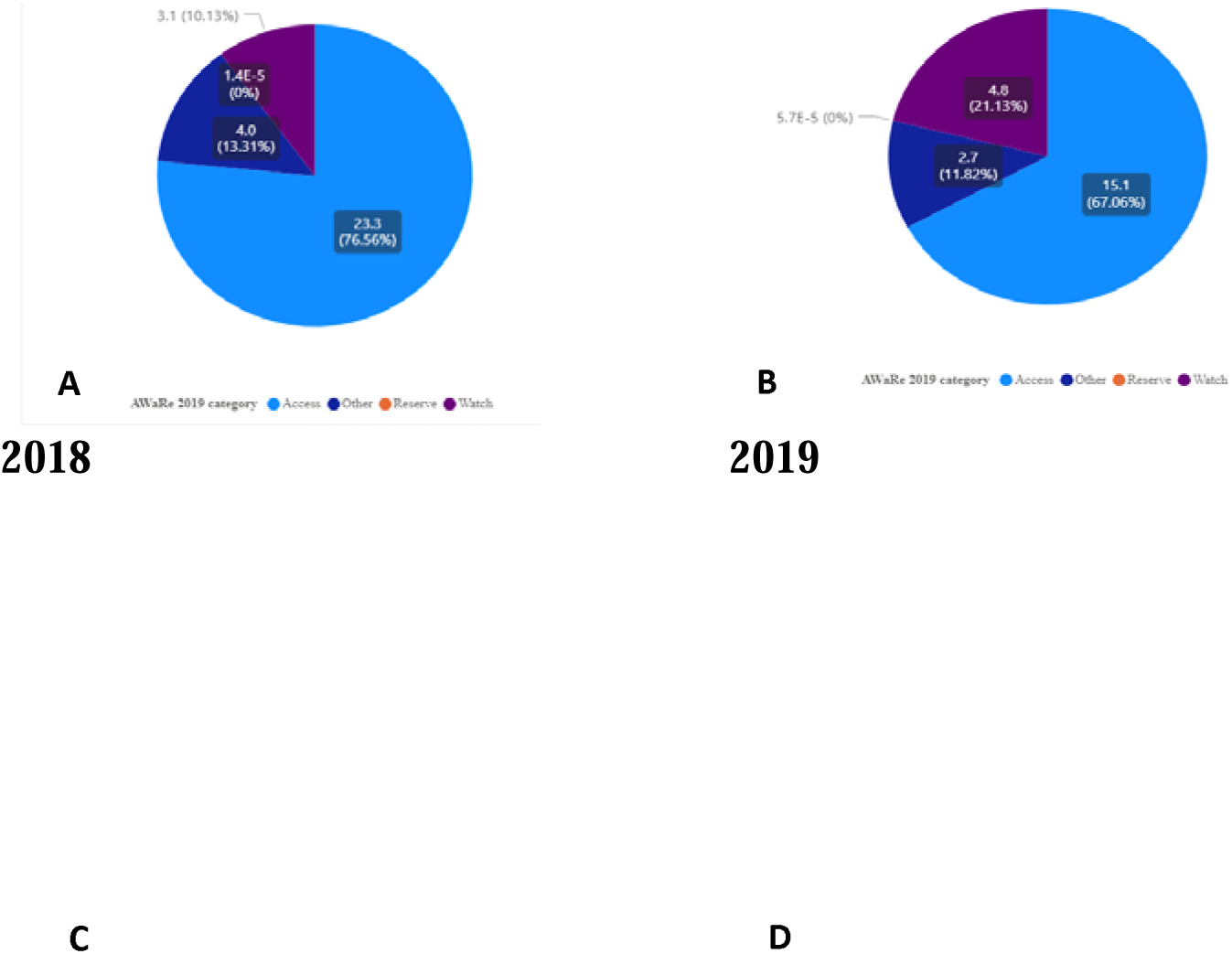
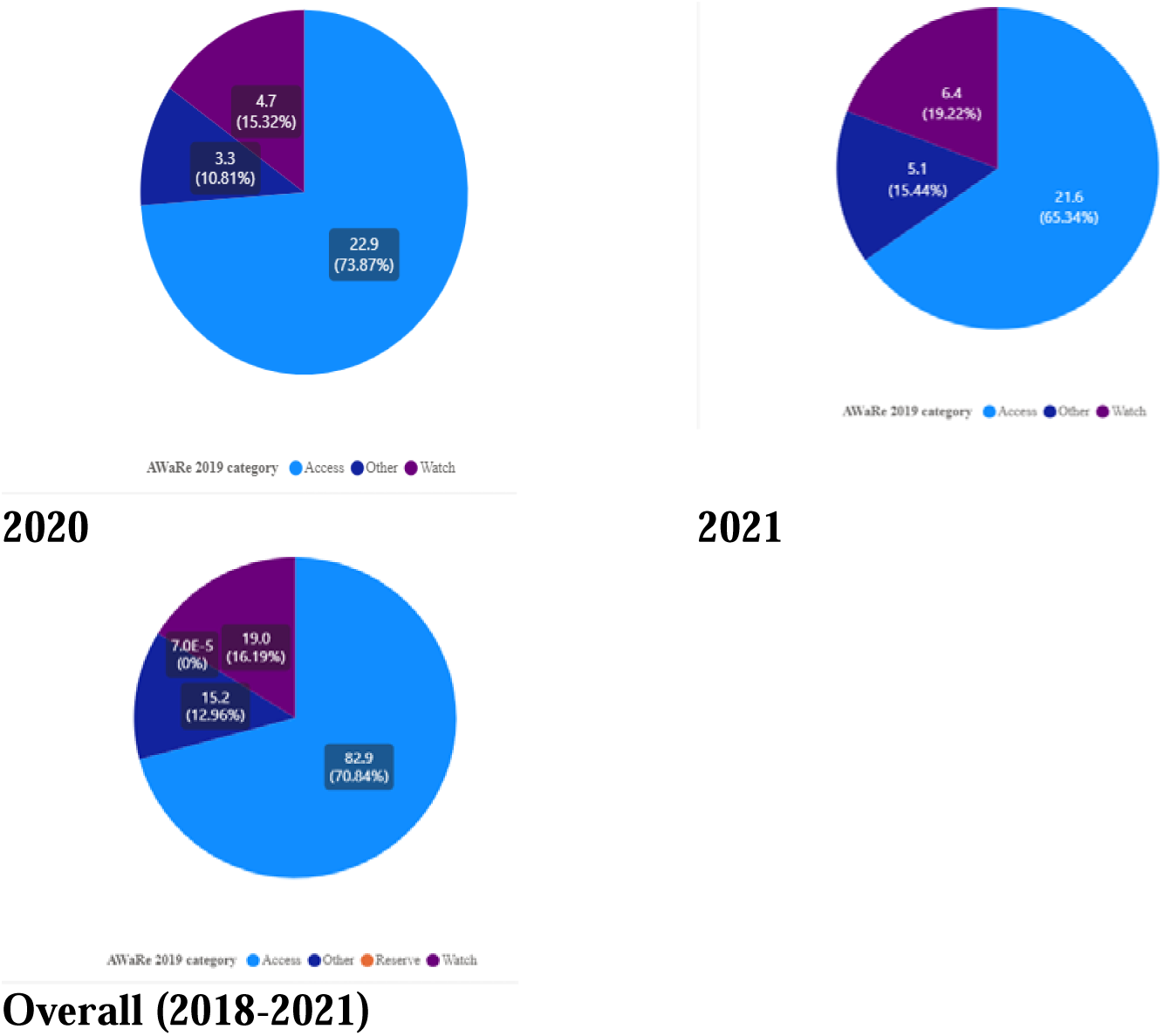
DID contribution per WHO AWaRe classification of antibiotics consumption from 2018-2021 (Panel A-D) and overall for four years (panel E)

The consumption of antibiotics was typically dominated by the Watch class (Supplementary Figure 2).

Using a paired samples t-test, the mean (M) and standard deviation (SD) of antibiotics consumption in the pre-COVID period (M = 1.018, SD = 3.311) was significantly different from the post-COVID period (M = 1.232, SD = 3.796), t (51= -2.513, p-value = 0.015 and paired sample correlation of 0.994 with effect size, as measured by Cohen’s d, being 0.312.

Overall, there was a 21% increase in the utilisation of antibiotics post-COVID-19. Azithromycin (J01FA10) increased by 163% during the pandemic (Table 2) and Supplementary Table 2.

**Table 2:** Percentage changes in consumption during COVID-19 for top 20 consumed antibiotics aggregated per level 5 WHO ATC classification in DID.

| Antibiotic (ATC Classification level 5) | Pre-COVID | Post-COVID | Total | % change |
| --- | --- | --- | --- | --- |
| Sulfamethoxazole + Trimethoprim (J01EE01) | 21.90206 | 24.134214 | 46.036274 | 10.2 |
| Amoxicillin (J01CA04) | 8.958047 | 12.109729 | 21.067776 | 35.2 |
| Ampicillin + Cloxacillin (J01CR50) | 4.957764 | 6.056899 | 11.014663 | 22.2 |
| Ciprofloxacin (J01MA02) | 3.320329 | 3.875245 | 7.195574 | 16.7 |
| Metronidazole (J01XD01) | 2.550591 | 3.007772 | 5.558363 | 17.9 |
| Azithromycin (J01FA10) | 1.163502 | 3.063967 | 4.227469 | 163.3 |
| Phenoxymethyl Penicillin (J01CE02) | 1.839013 | 2.112984 | 3.951997 | 14.9 |
| Erythromycin (J01FA01) | 1.103971 | 1.837235 | 2.941206 | 66.4 |
| Tinidazole (J01XD02) | 0.988315 | 1.399698 | 2.388013 | 41.6 |
| Amoxicillin + Clavulanate (J01CR02) | 0.81814 | 1.280495 | 2.098635 | 56.5 |
| Ceftriaxone (J01DD04) | 0.912814 | 1.147269 | 2.060083 | 25.7 |
| Cefalexin (J01DB01) | 0.956694 | 0.637602 | 1.594296 | -33.4 |
| Norfloxacin (J01MA06) | 0.929865 | 0.582542 | 1.512407 | -37.4 |
| Ampicillin (J01CA01) | 0.737038 | 0.699213 | 1.436251 | -5.1 |
| Gentamycin (J01GB03) | 0.184052 | 0.560144 | 0.744196 | 204.3 |
| Amoxicillin + Flucloxacillin (J01CR50) | 0.1963 | 0.216417 | 0.412717 | 10.2 |
| Ciprofloxacin + Tinidazole (J01RA11) | 0.219978 | 0.157204 | 0.377182 | -28.5 |
| Nitrofurantoin (J01XE01) | 0.220675 | 0.152053 | 0.372728 | -31.1 |
| Tetracycline (J01AA07) | 0.09207 | 0.222064 | 0.314134 | 141.2 |
| Chloramphenicol (J01BA01) | 0.210731 | 0.074664 | 0.285395 | -64.6 |

Considering level 3 of the ATC classification, we noted that antibiotics consumption varies by specific categories. Notably, beta-lactam antibacterials and penicillins (J01C) registered a significant 28.32% increase in consumption. A 4.96 DID increase between post-covid and pre-covid was noted for the beta-lactam antibacterials (Figure 4) and (Supplementary Table 3).

**Figure 4:**
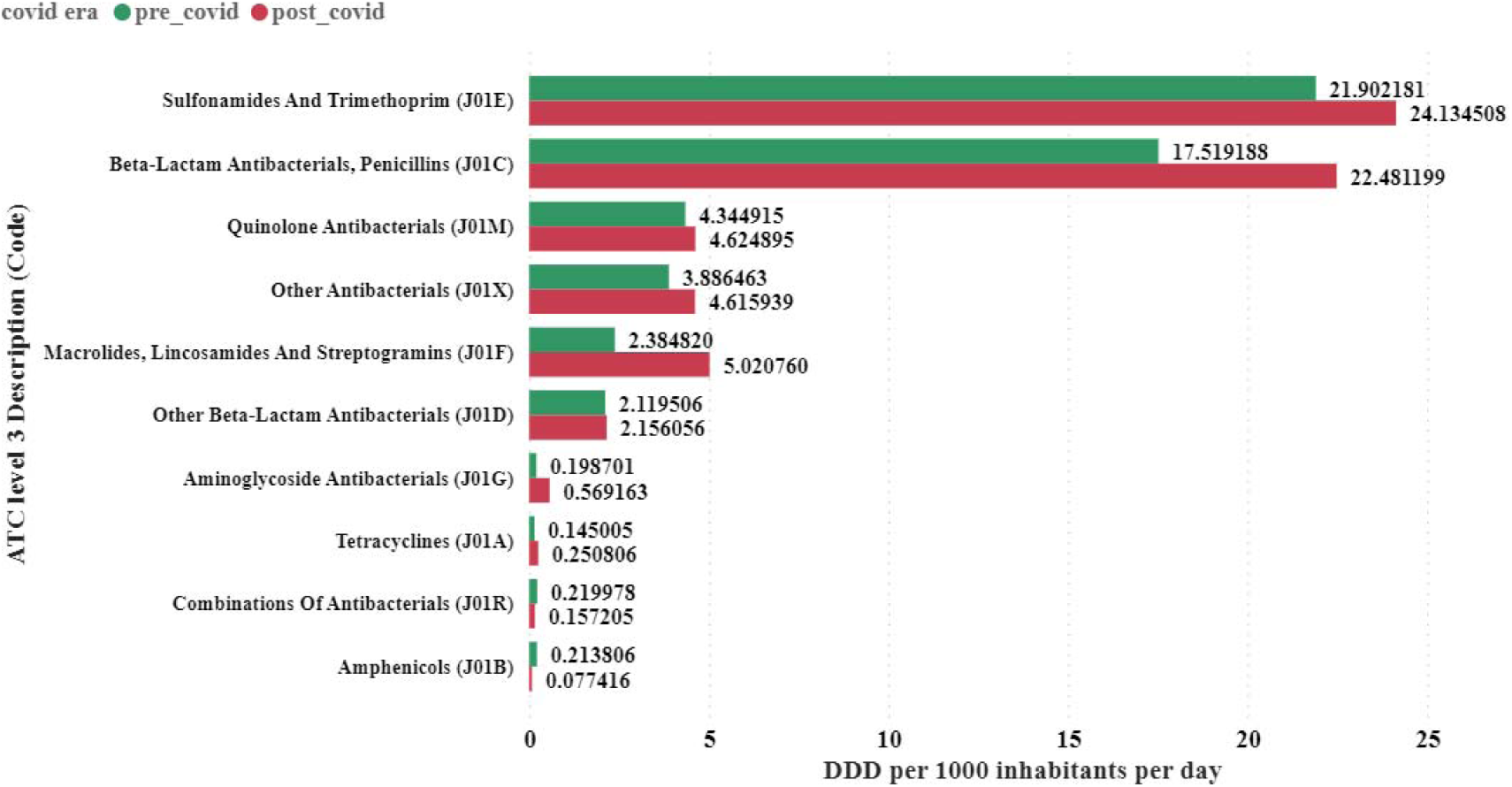
Contribution of each class (level 3 ATC classification) of antibiotics utilised in Tanzania from 2018 to 2021.

On the other hand, aminoglycoside antibacterials (J01G) exhibited a remarkable 186.44% increase. In contrast, amphenicols (J01B) experienced a substantial decrease –(63.79%). The class of macrolides, lincosamides and streptogramins (J01F) flagged a remarkable increase of 110.53% in consumption (Supplementary Table 3). In addition, annual trends of antibiotics at class 3 of the ATC classification were observed (Supplementary Table 4). Similar trends are indicated when considering level 4 ATC classification (Supplementary Table 5). It was noted that the level 3 ATC class of sulfonamides and trimethoprim (J01E) comprised 20.62% of all DID utilised, with the highest totals in pre- and post-COVID eras (Supplementary Table 4).

Overall, the consumption did not follow a linear pattern when fitting a linear curve (Figure 5A). In addition, the autoregressive integrated moving average (ARIMA) (0, 1, 0) model (Figure 5B) predicts a significant increase in utilisation and forecasts the trends of antibiotics up to the period 2027 modelled using the utilisation trend between 2018 and 2021.

**Figure 5:**
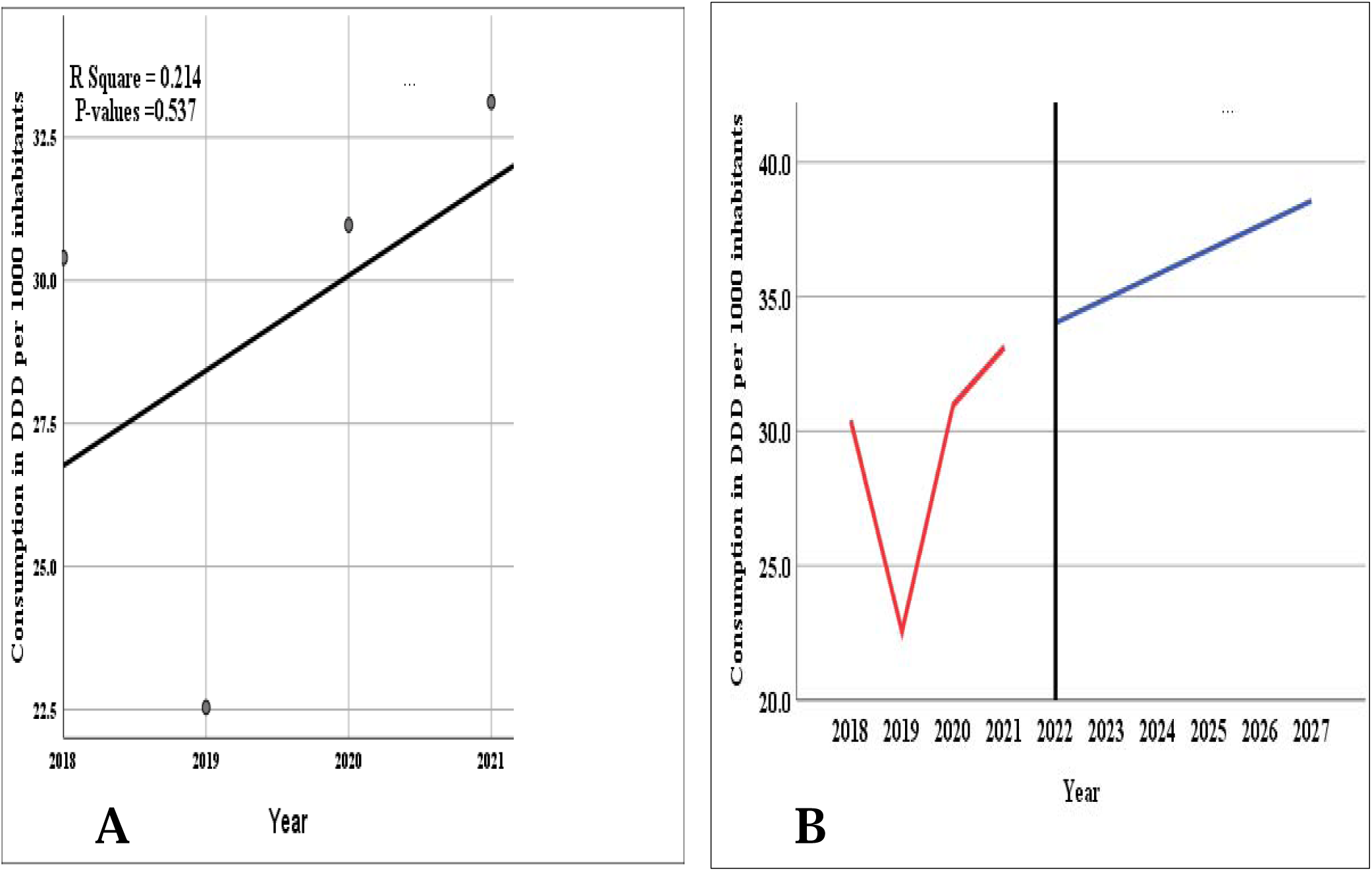
A) Linear curve estimation showing trends of total consumed antibiotics over four years from 2018 to 2021 B) ARIMA model prediction of consumption from 2018 through 2027.

The model estimates that by 2022 and 2027, the DID will reach 34.03 and 38.56 DIDs, respectively.

The top 15 importers of these antibiotics contributed to 90.06% of all DIDs, with the Medical Stores Department (MSD) leading by contributing 20.27% (Figure 6) and (Supplementary Table 6).

**Figure 6.**
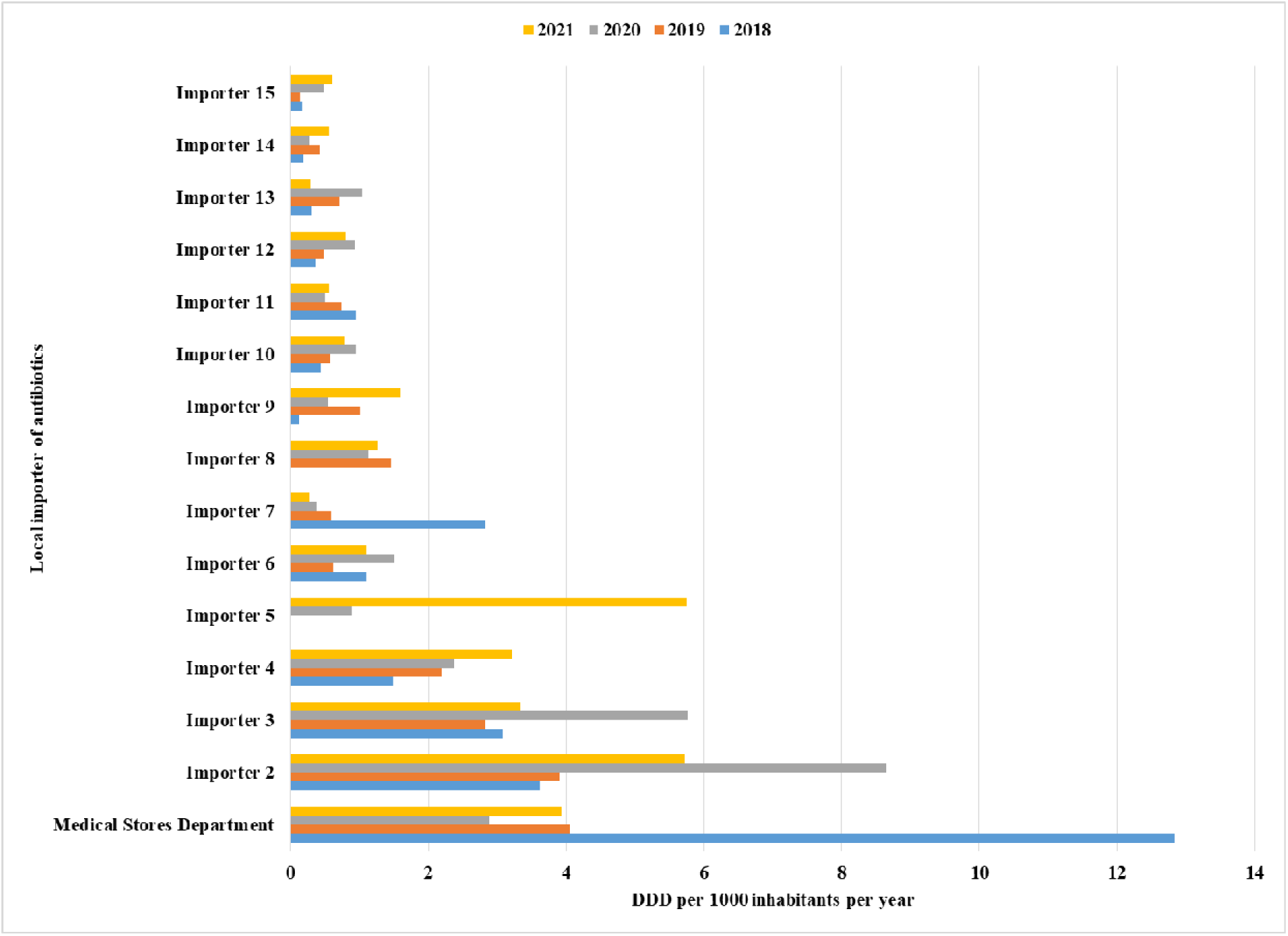
Top 15 local importers of antibiotics utilised in Tanzania between 2018 and 2021.

Regarding suppliers, the top 15 contributed 70.78% of the DID of antibiotics in Tanzania (Figure 7) and (Supplementary Table 7). The suppliers were from China, India and Kenya, consistent with the map distribution (Figure 1).

**Figure 7:**
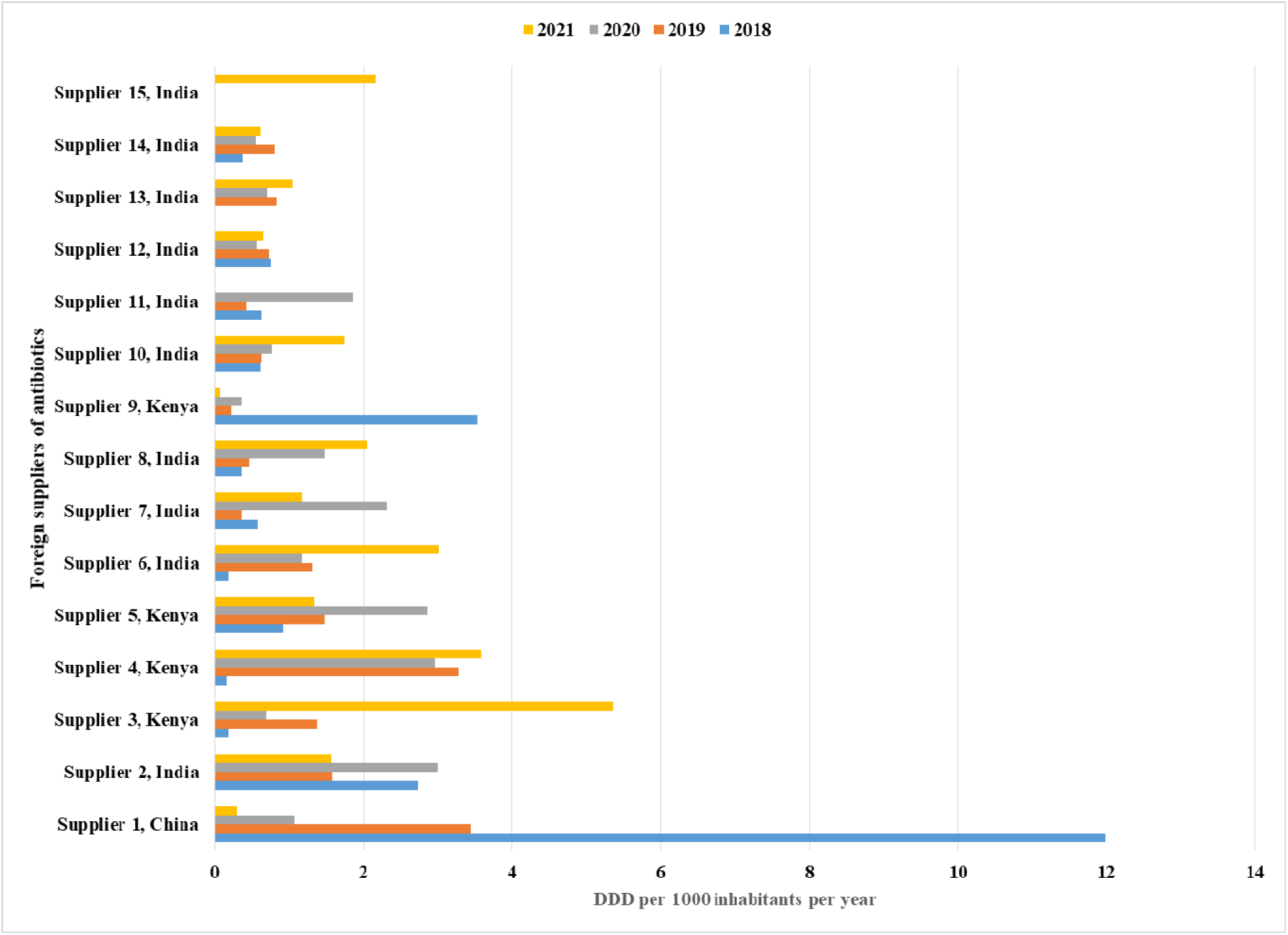
Top 15 foreign suppliers of antibiotics utilised in Tanzania between 2018 and 2021.

## Discussion

The Global Action Plan for Antimicrobial Resistance aims to address the mounting challenge of increasing antimicrobial resistance (AMR) through the surveillance of antimicrobial use (AMU) and the development of antimicrobial stewardship (AMS) programs. Consequently, in Tanzania, AMS was introduced through the NAP on AMR^16^ in 2017 and the second version of NAP^17^ 2023-2028 focuses on monitoring AMU in humans and animals. ^15^

In this study, we observed an annual increase in the total consumption of antibiotics, reaching 117.02 DDI over four years. The consumption was 64.09 DDI in the post-COVID era and 52.93 DDI in the pre-COVID era. Nevertheless, the mean is 29.25 (±4.01) compared to the mean of 22.07 (±48.85) DID in 2010 to 2016 consumption data in Tanzania^15^.

A paired samples t-test indicated a significant difference in means 1.018 vs. = 1.232 in pre and post-COVID periods, respectively, with p-value = 0.015 and paired sample correlation of 0.994. This result suggests a statistically significant increase in antibiotics consumption during the pandemic. The effect size, as measured by Cohen’s d, was 0.312, suggesting a small but practically significant increase. The high correlation between pre-COVID and post-COVID consumption (r = 0.994) reinforces the reliability of these findings, suggesting that the COVID-19 pandemic had a notable impact on antibiotics consumption in Tanzania.

The combined usage of all antibiotics increased by 21.1% from the pre-COVID period to the post-COVID period. An increase was noted for gentamycin (J01GB03) at +204.3%, followed by azithromycin (J01FA10) at +163.3% and tetracycline (J01AA07) at +141.2%. A decrease was observed in chloramphenicol (J01BA01) (-64.6%), norfloxacin (J01MA06) (-37.4%), and nitrofurantoin (J01XE01) (-31.1%). A % increase in azithromycin was noted in other studies in LMICS and HICs. ^19^ A study in Croatia showed that azithromycin distribution increased from 1.76 in 2017 to 2.01 Days of Therapy (DOTS) units/1000 inhabitant-days in 2017–2020, indicating azithromycin overuse.^20^ Other reports during the pandemic showed that azithromycin consumption increased up to 3 times compared to the pre-COVID period.^19–21^

Interestingly, the popularity of azithromycin emerged from reports of its antiviral activity and also from early pandemic reports of screening indicating potential activity for SARS-CoV-2 alone or in combination with hydroxychloroquine.^22^ Later, several randomised clinical trials (RCTs) suggested that azithromycin does not reduce hospital admissions, respiratory failure, or death when compared to conventional therapy, and therefore, azithromycin should no longer be used to treat COVID-19.^23–28^

Several studies have revealed a significant increase in resistance to azithromycin in some strains of *Neisseria gonorrhoeae*.^22,29^ Antibiotic resistance against azithromycin also increased in *E. coli*^30^ and *Streptococcus pneumoniae.*^31^ Therefore, continued use of azithromycin should have been limited to infections for which azithromycin is recommended rather than COVID-19. ^22^

Examining the consumption at level 3 of ATC classification, we noted a remarkable increase during the post-COVID-19 era of beta-lactam antibacterials, which penicillins (J01C) and aminoglycoside antibacterials (J01G) exhibited. At the same time, amphenicols (J01B) experienced a substantial decrease up to -63.79%. The use of macrolides, lincosamides and streptogramins (J01F) also increased remarkably by 110.53%. This finding underscores the specific impact of the pandemic on the consumption of these antibiotics; the major contributor to this increase was azithromycin.

The overall consumption of antibiotics increased from 52.935 DID (pre-COVID) to 64.088 DID (post-COVID), with a total change of 21.07%.

Overall, the ATC level 3 class of sulfonamides and trimethoprim (J01E) ranked the top consumed group with only 10.19% increase in consumption, suggesting continued reliance on this class of antibiotics during the pandemic. This could be due to their effectiveness against certain infections and wide availability, especially for HIV/AIDS patients. This is usually indicated by the higher contribution of Sulfamethoxazole + Trimethoprim (J01EE01) used in the HIV program.

For tetracycline (level 3 class J01A), there was a moderate consumption increase from 0.145 DID (pre-COVID) to 0.251 DID (post-COVID), a 72.96% increase, even though this class ranked lower compared to previous studies in Tanzania where the class was among the top contributors of consumed antibiotics.^15^ It is important to note that the percentage change in usage should be taken with caution since it is calculated based on a relatively small difference in values, and the absolute values of DID for each antibiotic may vary significantly.

A recent study conducted in Cameroon during the COVID-19 pandemic revealed that antibiotics were highly overused and misused, leading to increased AMR.^32^

This study is one of the few conducted in sub-Saharan Africa to estimate antibiotics utilisation at the national level. The data indicate an increase in the consumption of antibiotics during the pandemic, with a mean of 29.25 DIDs utilised in Tanzania between 2018 and 2021. This average was less than that studied between 2010 and 2016 in Tanzania, where the mean was 57.4 DIDs over seven years. The study conducted in Tanzania from 2017 to 2019 also reported a slightly higher average compared to this study.^33^

These results highlight the importance of expanding the monitoring of AMU and implementing AMS programs to address the issue of AMR, especially during global health crises such as the COVID-19 pandemic. The observed changes in antibiotic consumption highlight the need for continued monitoring and the development of interventions to ensure the rational use of antibiotics since the increase in overall consumption may contribute to AMR. Antibiotic stewardship programs must be emphasised post-COVID in the healthcare landscape.

The ARIMA forecasted that antibiotics consumption would increase between 2022 and 2027 to reach 34.03 and 38.56 DIDs, respectively. This value is less than the value predicted with the data from 2010–2016 which estimated that by 2022, the total of antibiotics consumed would reach 89.60 DIDs.^15^ This may reflect the impact of AMS under the NAP implementation. ^16,34,35^

## Limitations of the study

This study has some limitations. First, it relies on data from importation records of NMRA, potentially subject to variability and errors, which could impact the accuracy of the results. The data does not account for imported antibiotics that had expired or were re-exported to neighbouring countries. These records also exclude efforts by manufacturers that produced some antibiotics locally during the pandemic. Second, while it explores the association between the pandemic and antibiotic consumption, it does not establish causality, as other unaccounted factors may influence consumption trends. Third, the study’s aggregated data may not capture regional variations, which is important for understanding local healthcare practices and policies. Additionally, the analysis’s specific timeframe during the pandemic may not account for evolving healthcare practices and AMS interventions, and the study does not delve into the underlying factors driving these trends. Further research is warranted to understand the factors driving these consumption trends and their potential impact on public health. Results need to be interpreted with these limitations in mind.

## Conclusion

This study highlights an increase in the consumption of antibiotics during the COVID-19 pandemic, indicating that the COVID-19 pandemic impacted the rise in antibiotics consumption in Tanzania.

## Recommendations

The surge in AMU post-COVID-19 calls for continuous monitoring and interventions to promote rational use of antibiotics to combat AMR through AMS and ensure long-term effectiveness in managing infectious diseases. In addition, more studies using data from community pharmacies and hospitals are needed for a more accurate representation of patient antibiotic consumption.

## Availability of Data and Materials

All data are included in this article.

## Acknowledgement

We thank the Tanzania Medicines and Medical Devices Authority (TMDA) staff for providing the importation data.

## Funding

This work did not receive any specific funding.

## Transparency declarations

None to declare

## Supplementary data

Figures and Tables are available as Supplementary data at JAC-AMR Online.

---

**Supplementary Figure 1:**
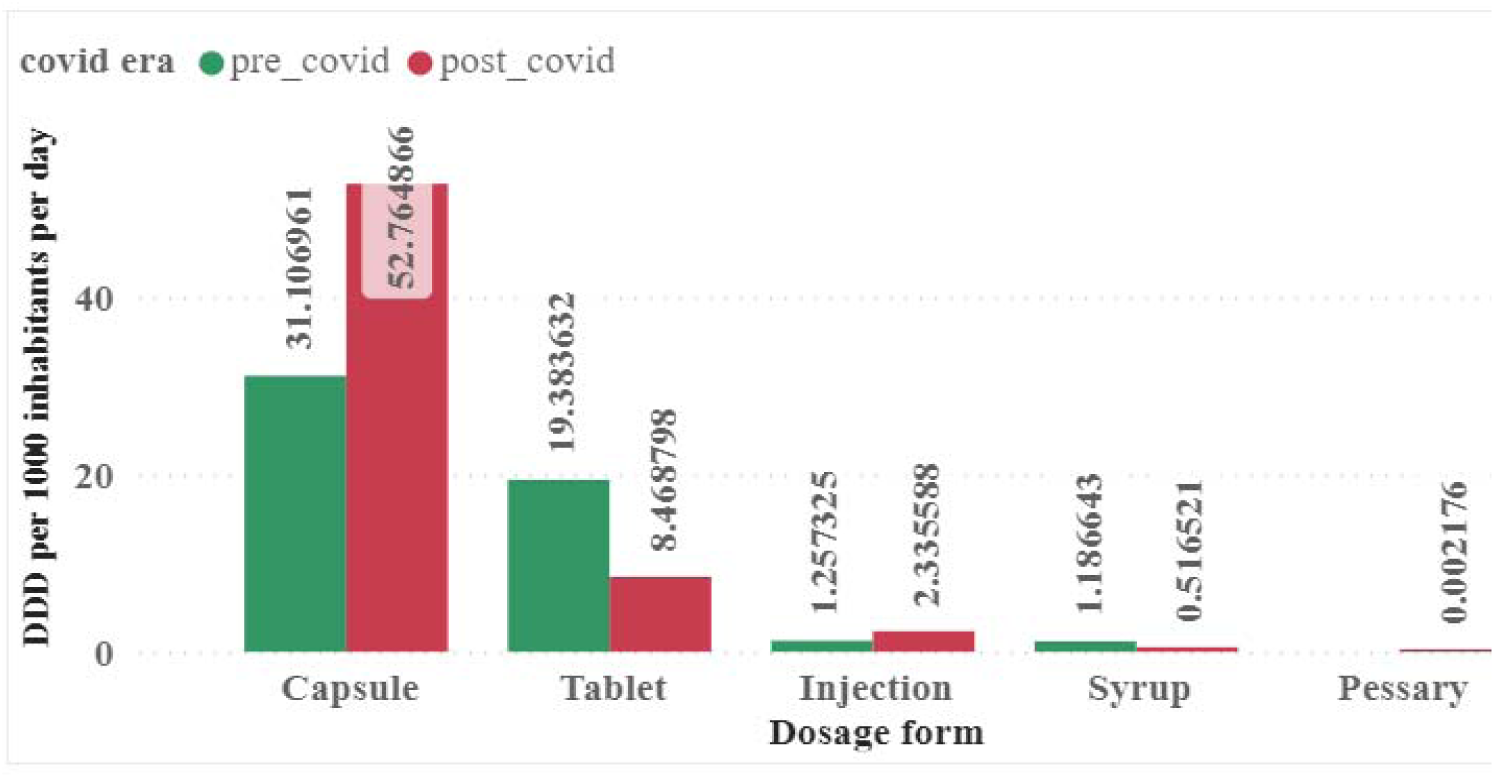
Contribution of antibiotics consumption per dosage and by COVID-19 era.

**Supplementary Figure 2:**
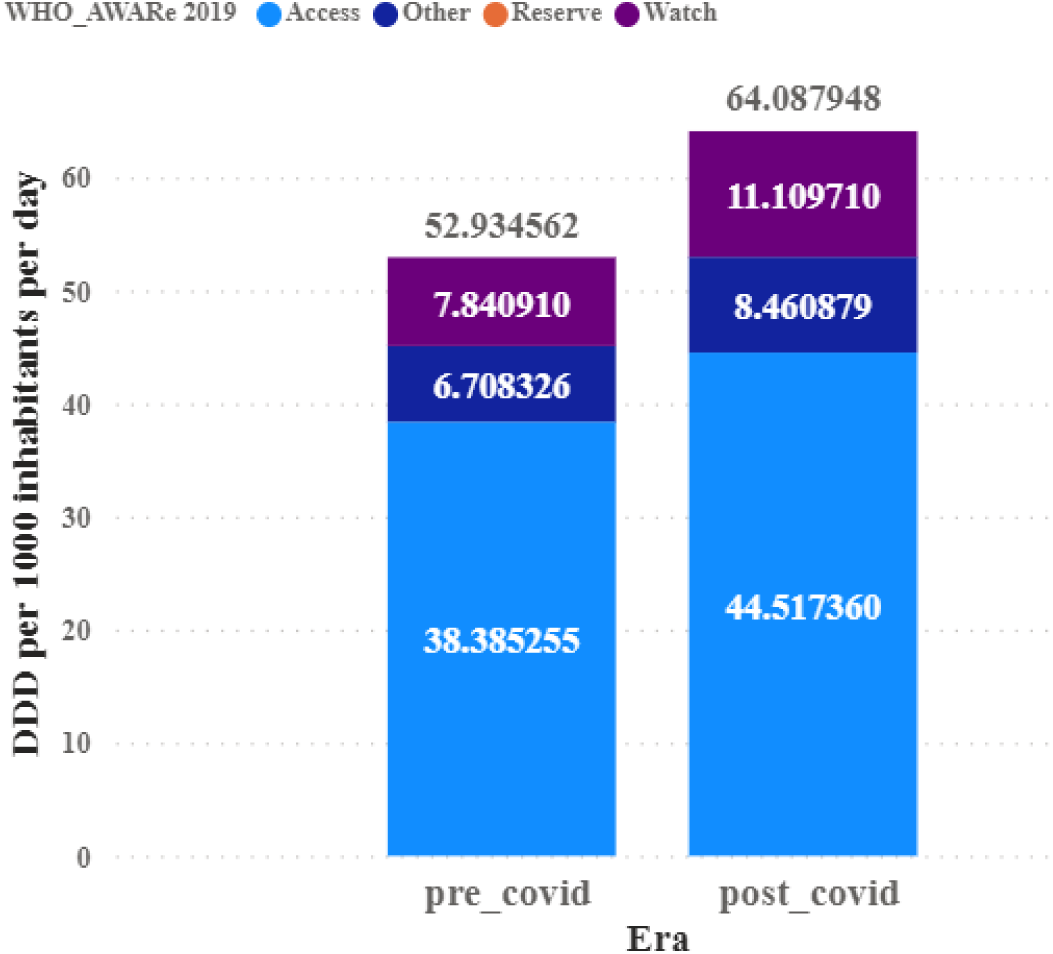
Distribution of Defined Daily Dose (DDD per 1000 inhabitants per day (DID)) of antibiotics per the World Health Organization’s AWaRe class for antibiotics utilised in Tanzania from 2018 to 2020.

**Supplementary Table 1:** Annual contribution of consumption of antibiotics per dosage form.

|  | Year | % contribution |  |  |  |
| --- | --- | --- | --- | --- | --- |
| Dosage form | 2018 | 2019 | 2020 | 2021 | All time |
| Capsules | 52.5 | 67.2 | 84.7 | 80.2 | 71.1 |
| Injections | 2.3 | 2.5 | 3.9 | 3.4 | 3.0 |
| Pessaries |  |  |  | 0.0 | 0.0 |
| Syrup | 2.7 | 1.6 | 0.7 | 0.9 | 1.5 |
| Tablets | 42.5 | 28.7 | 10.8 | 15.5 | 24.4 |
| **Total** | **100.0** | **100.0** | **100.0** | **100.0** | 100.0 |

**Supplementary Table 2:** Percentage changes in consumption during COVID-19 for top 20 consumed antibiotics aggregated per level 5 WHO ATC classification in DID.

| Antibiotic (ATC level 5 code) | DID |  |  | % Change |
| --- | --- | --- | --- | --- |
|  | Pre COVID | Post COVID | Total |  |
| Sulfamethoxazole + Trimethoprim (J01EE01) | 21.90206 | 24.134214 | 46.036274 | 10.2 |
| Amoxicillin (J01CA04) | 8.958047 | 12.109729 | 21.067776 | 35.2 |
| Ampicillin + Cloxacillin (J01CR50) | 4.957764 | 6.056899 | 11.014663 | 22.2 |
| Ciprofloxacin (J01MA02) | 3.320329 | 3.875245 | 7.195574 | 16.7 |
| Metronidazole (J01XD01) | 2.550591 | 3.007772 | 5.558363 | 17.9 |
| Azithromycin (J01FA10) | 1.163502 | 3.063967 | 4.227469 | 163.3 |
| Phenoxymethyl Penicillin (J01CE02) | 1.839013 | 2.112984 | 3.951997 | 14.9 |
| Erythromycin (J01FA01) | 1.103971 | 1.837235 | 2.941206 | 66.4 |
| Tinidazole (J01XD02) | 0.988315 | 1.399698 | 2.388013 | 41.6 |
| Amoxicillin + Clavulanate (J01CR02) | 0.81814 | 1.280495 | 2.098635 | 56.5 |
| Ceftriaxone (J01DD04) | 0.912814 | 1.147269 | 2.060083 | 25.7 |
| Cefalexin (J01DB01) | 0.956694 | 0.637602 | 1.594296 | -33.4 |
| Norfloxacin (J01MA06) | 0.929865 | 0.582542 | 1.512407 | -37.4 |
| Ampicillin (J01CA01) | 0.737038 | 0.699213 | 1.436251 | -5.1 |
| Gentamycin (J01GB03) | 0.184052 | 0.560144 | 0.744196 | 204.3 |
| Amoxicillin + Flucloxacillin (J01CR50) | 0.1963 | 0.216417 | 0.412717 | 10.2 |
| Ciprofloxacin + Tinidazole (J01RA11) | 0.219978 | 0.157204 | 0.377182 | -28.5 |
| Nitrofurantoin (J01XE01) | 0.220675 | 0.152053 | 0.372728 | -31.1 |
| Tetracycline (J01AA07) | 0.09207 | 0.222064 | 0.314134 | 141.2 |
| Chloramphenicol (J01BA01) | 0.210731 | 0.074664 | 0.285395 | -64.6 |
| Cefixime (J01DD08) | 0.089679 | 0.18745 | 0.277129 | 109.0 |
| Clarithromycin (J01FA09) | 0.116368 | 0.116796 | 0.233164 | 0.4 |
| Ornidazole (J01XD03) | 0.124153 | 0.053845 | 0.177998 | -56.6 |
| Cefuroxime (J01DC02) | 0.073682 | 0.04626 | 0.119942 | -37.2 |
| Ofloxacin (J01MA01) | 0.052129 | 0.06318 | 0.115309 | 21.2 |
| Cefadroxil (J01DB05) | 0.041038 | 0.050992 | 0.09203 | 24.3 |
| Levofloxacin (J01MA13) | 0.008836 | 0.082444 | 0.09128 | 833.0 |
| Doxycycline (J01AA02) | 0.052935 | 0.028352 | 0.081287 | -46.4 |
| Cefotaxime (J01DD01) | 0.005643 | 0.066155 | 0.071798 | 1072.3 |
| Cefpodoxime (J01DD13) | 0.02914 | 0.006786 | 0.035926 | -76.7 |
| Moxifloxacin (J01MA15) | 0.015046 | 0.012868 | 0.027914 | -14.5 |
| Dexamethasone + Neomycin + Polymyxin B (J01GB05) | 0.014225 | 0.008568 | 0.022793 | -39.8 |
| Lomefloxacin (J01MA07) | 0.010553 | 0.008616 | 0.019169 | -18.4 |
| Meropenem (J01DH02) | 0.00761 | 0.009849 | 0.017459 | 29.4 |
| Ampicillin + enzyme inhibitor (J01CA51) | 0.011029 | 0.004753 | 0.015782 | -56.9 |
| Nalidixic Acid (J01MB02) | 0.008157 |  | 0.008157 | -100.0 |
| Lignocaine + Chloramphenicol + Beclomethasone Dipropionate + Clotrimazole (J01BA01) | 0.003076 | 0.002752 | 0.005828 | -10.5 |
| Clindamycin (J01FF01) | 0.00078 | 0.002762 | 0.003542 | 254.1 |
| Spectinomycin (J01XX04) | 0.001507 | 0.00086 | 0.002367 | -42.9 |
| Cefepime (J01DE01) | 0.001017 | 0.000937 | 0.001954 | -7.9 |
| Flucloxacillin (J01CF05) | 0.001857 |  | 0.001857 | -100.0 |
| Vancomycin (J01XA01) | 0.000392 | 0.001189 | 0.001581 | 203.3 |
| Cefoperazone + combinations (J01DD62) | 0.000996 | 0.000461 | 0.001457 | -53.7 |
| Cefoperazone + Sulbactam (J01DD62) | 0.000497 | 0.000615 | 0.001112 | 23.7 |
| Bacitracin + Neomycin + Polymyxin B (J01XX10 ) | 0.000561 | 0.000423 | 0.000984 | -24.6 |
| Cefpirome (J01DE02) |  | 0.000867 | 0.000867 | NA |
| Cilastatin + Imipenem (J01DH51) | 0.0003 | 0.000487 | 0.000787 | 62.3 |
| Cloxacillin (J01CF02) |  | 0.000709 | 0.000709 | NA |
| Amikacin (J01GB06) | 0.00027 | 0.000181 | 0.000451 | -33.0 |
| Cefazolin (J01DB04) | 0.000174 | 0.000275 | 0.000449 | 58.0 |
| Tylosin Tartrate + Doxycycline Hyclate (J01AA02) |  | 0.000389 | 0.000389 | NA |
| PolyMyxin B (J01XB02) | 0.000199 | 0.0001 | 0.000299 | -49.7 |
| Neomycin (J01GB05) |  | 0.000267 | 0.000267 | NA |
| Roxithromycin (J01FA06) | 0.000199 |  | 0.000199 | -100.0 |
| Sulfadiazine + Trimethoprim (J01EE02) | 0.000051 | 0.000136 | 0.000187 | 166.7 |
| Kanamycin (J01GB04) | 0.000153 |  | 0.000153 | -100.0 |
| Sulfadimidine (J01EB03) |  | 0.000123 | 0.000123 | NA |
| Trimethoprim (J01EA01) | 0.00007 | 0.000035 | 0.000105 | -50.0 |
| Cefaclor (J01DC04) | 0.000077 | 0.000027 | 0.000104 | -64.9 |
| Imipenem + enzyme inhibitor (J01DH56) | 0.000089 |  | 0.000089 | -100.0 |
| Ceftazidime (J01DD02) | 0.000056 | 0.000024 | 0.00008 | -57.1 |
| Linezolid (J01XX08) | 0.00007 |  | 0.00007 | -100.0 |
| Tobramycin (J01GB01) | 0.000001 | 0.000002 | 0.000003 | 100.0 |
| Isoniazid + Pyridoxine + Sulfamethoxazole + Trimethoprim (J04AM08) |  | 0.000001 | 0.000001 | NA |
| Ampicillin + Sulbactam (J01CR01) | 0 |  | 0 | NA |
| Azithromycin + fluconazole + secnidazole (J01RA07) |  | 0 | 0 | NA |
| Erythromycin + combinations (J01FA01) |  | 0 | 0 | N.A. |
| **Period Total** | **52.934564** | **64.087946** | **117.02251** | 21.1 |

**Supplementary Table 3:**
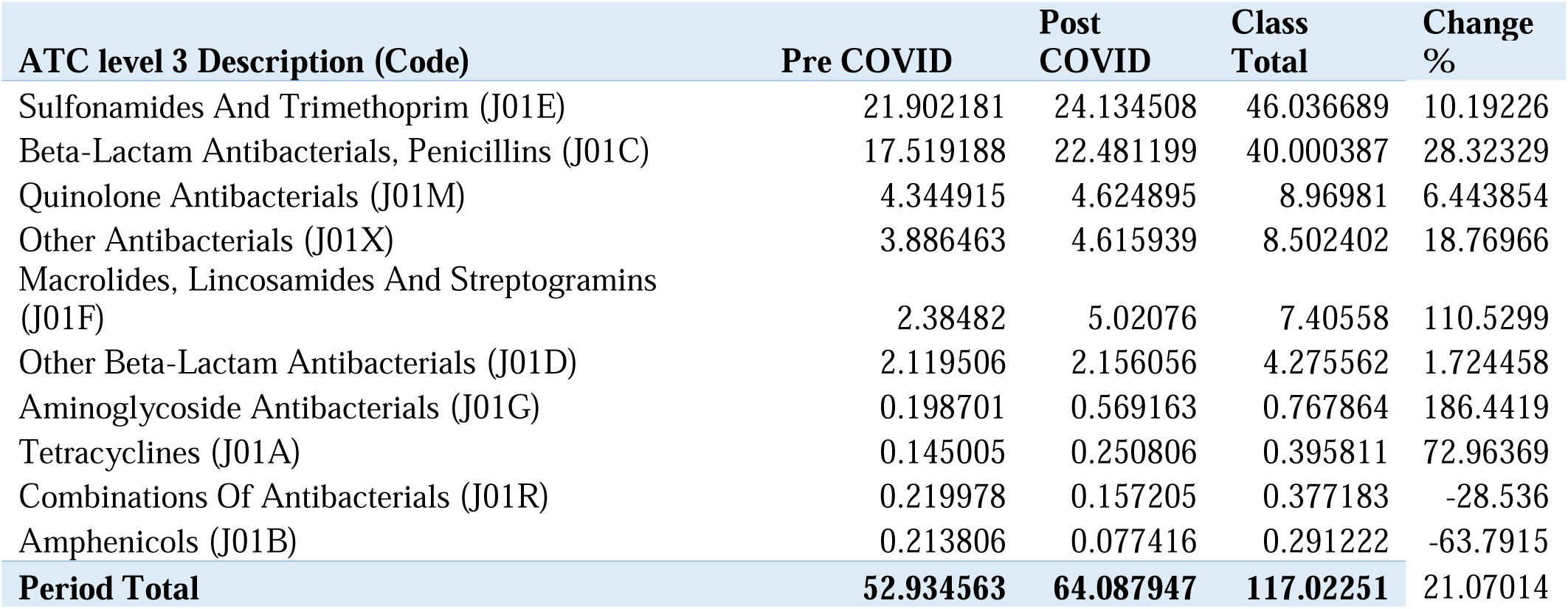
Consumption aggregated at ATC level 3 in the pre-COVID-19 and post-COVID-19 era in Tanzania.

**Supplementary Table 4:** Consumption aggregated at ATC level 3 from 2018 to 2021 in Tanzania.

| ATC Class level 3 | 2018 | 2019 | 2020 | 2021 | Four Years Total |
| --- | --- | --- | --- | --- | --- |
| Sulfonamides And Trimethoprim (J01E) | 14.617083 | 7.285097 | 12.50324 | 11.631268 | 46.036688 |
| Beta-Lactam Antibacterials, Penicillins (J01C) | 10.173568 | 7.34562 | 11.12656 | 11.35464 | 40.000388 |
| Quinolone Antibacterials (J01M) | 1.462383 | 2.882532 | 2.309063 | 2.315832 | 8.96981 |
| Other Antibacterials (J01X) | 1.784649 | 2.101814 | 1.960377 | 2.655562 | 8.502402 |
| Macrolides, Lincosamides And Streptogramins (J01F) | 1.064215 | 1.320605 | 1.65612 | 3.364641 | 7.405581 |
| Other Beta-Lactam Antibacterials (J01D) | 0.872695 | 1.24681 | 1.157145 | 0.998911 | 4.275561 |
| Aminoglycoside Antibacterials (J01G) | 0.155795 | 0.042906 | 0.07892 | 0.490243 | 0.767864 |
| Tetracyclines (J01A) | 0.042675 | 0.10233 | 0.10106 | 0.149746 | 0.395811 |
| Combinations Of Antibacterials (J01R) | 0.060057 | 0.159921 | 0.024369 | 0.132836 | 0.377183 |
| Amphenicols (J01B) | 0.165188 | 0.048619 | 0.051204 | 0.026212 | 0.291223 |
| Drugs For Treatment Of Tuberculosis (J04A) |  |  |  | 0.000001 | 0.000001 |
| **Year Total** | **30.398308** | **22.536254** | **30.968058** | **33.119892** | **117.022512** |

**Supplementary Table 5:** Consumption aggregated at ATC level 4 from 2018 to 2021 in Tanzania.

| ATC Class level 4 | 2018 | 2019 | 2020 | 2021 | Four years Total |
| --- | --- | --- | --- | --- | --- |
| Amphenicols (J01BA) | 0.165188 | 0.048619 | 0.051204 | 0.026212 | 0.291223 |
| Beta-Lactamase Resistant Penicillins (J01CF) | 0.000632 | 0.001225 | 0.000709 |  | 0.002566 |
| Beta-Lactamase Sensitive Penicillins (J01CE) | 0.97563 | 0.863383 | 1.066293 | 1.04669 | 3.951996 |
| Carbapenems (J01DH) | 0.006551 | 0.001447 | 0.00222 | 0.008116 | 0.018334 |
| Combinations Of Antibacterials (J01RA) | 0.060057 | 0.159921 | 0.024369 | 0.132836 | 0.377183 |
| Combinations Of Drugs For Treatment Of Tuberculosis (J04AM) |  |  |  | 0.000001 | 0.000001 |
| Combinations Of Penicillins, Incl. Beta-Lactamase Inhibitors (J01CR) | 3.626861 | 2.345343 | 3.431805 | 4.122006 | 13.526015 |
| Combinations Of Sulphonamides And Trimethoprim Incl. Derivatives (J01EE) | 14.617026 | 7.285085 | 12.503105 | 11.631245 | 46.036461 |
| First Generation Cephalosporins (J01DB) | 0.310298 | 0.687607 | 0.376731 | 0.312137 | 1.686773 |
| Fluoroquinolones (J01MA) | 1.454226 | 2.882532 | 2.309063 | 2.315832 | 8.961653 |
| Fourth Generation Cephalosporins (J01DE) | 0.000656 | 0.000361 | 0.00063 | 0.001174 | 0.002821 |
| Glycopeptide Antibacterials (J01XA) | 0.000293 | 0.000098 | 0.000166 | 0.001022 | 0.001579 |
| Imidazole Derivatives (J01XD) | 1.710669 | 1.952389 | 1.893326 | 2.567989 | 8.124373 |
| Lincosamides (J01FF) | 0.000276 | 0.000503 | 0.000896 | 0.001866 | 0.003541 |
| Macrolides (J01FA) | 1.063939 | 1.320101 | 1.655224 | 3.362775 | 7.402039 |
| Other Aminoglycosides (J01GB) | 0.155795 | 0.042906 | 0.07892 | 0.490243 | 0.767864 |
| Other Antibacterials (J01XX) | 0.001017 | 0.001122 | 0 | 0.001283 | 0.003422 |
| Other Quinolones (J01MB) | 0.008157 |  |  |  | 0.008157 |
| Penicillins With Extended Spectrum (J01CA) | 5.570445 | 4.135668 | 6.627752 | 6.185944 | 22.519809 |
| Second Generation Cephalosporins (J01DC) | 0.042137 | 0.031622 | 0.009737 | 0.03655 | 0.120046 |
| Short-Acting Sulfonamides (J01EB) |  |  | 0.000123 |  | 0.000123 |
| Tetracyclines (J01AA) | 0.042675 | 0.10233 | 0.10106 | 0.149746 | 0.395811 |
| Third Generation Cephalosporins (J01DD) | 0.513052 | 0.525773 | 0.767827 | 0.640932 | 2.447584 |
| (Not classified) | 0.072727 | 0.148216 | 0.066897 | 0.085291 | 0.373131 |
| **Year Total** | **30.398307** | **22.536251** | **30.968057** | **33.11989** | **117.022505** |

**Supplementary Table 6:** Top (local) importers of antibiotics utilised in Tanzania between 2018 and 2021, with the proportion of each importer in DID in four years and the cumulative contribution for each local importer.

| Importer | 2018 | 2019 | 2020 | 2021 | Total | Proportion | Cumulative % contribution |
| --- | --- | --- | --- | --- | --- | --- | --- |
| Medical Stores Department | 12.832301 | 4.068537 | 2.884632 | 3.929656 | 23.715126 | 20.26544022 | 20.2654402 |
| Importer 2 | 3.61511 | 3.910759 | 8.653665 | 5.725965 | 21.905499 | 18.71904794 | 38.9844882 |
| Importer 3 | 3.08651 | 2.834042 | 5.760543 | 3.326914 | 15.008009 | 12.82489114 | 51.8093793 |
| Importer 4 | 1.490486 | 2.196296 | 2.374687 | 3.217808 | 9.279277 | 7.929480676 | 59.73886 |
| Importer 5 |  |  | 0.880014 | 5.746847 | 6.626861 | 5.662894452 | 65.4017544 |
| Importer 6 | 1.105711 | 0.629544 | 1.501565 | 1.104843 | 4.341663 | 3.710109404 | 69.1118638 |
| Importer 7 | 2.82273 | 0.603366 | 0.374427 | 0.270447 | 4.07097 | 3.478792362 | 72.5906562 |
| Importer 8 |  | 1.462745 | 1.136031 | 1.264955 | 3.863731 | 3.301699077 | 75.8923553 |
| Importer 9 | 0.130747 | 1.015805 | 0.541305 | 1.593834 | 3.281691 | 2.804324666 | 78.6966799 |
| Importer 10 | 0.438368 | 0.577306 | 0.945605 | 0.792103 | 2.753382 | 2.352865354 | 81.0495453 |
| Importer 11 | 0.948832 | 0.737385 | 0.49991 | 0.553922 | 2.740049 | 2.341471819 | 83.3910171 |
| Importer 12 | 0.357483 | 0.482545 | 0.94044 | 0.807016 | 2.587484 | 2.211099462 | 85.6021166 |
| Importer 13 | 0.314455 | 0.718956 | 1.039605 | 0.300066 | 2.373082 | 2.027885132 | 87.6300017 |
| Importer 14 | 0.186325 | 0.427968 | 0.267492 | 0.554332 | 1.436117 | 1.227214362 | 88.8572161 |
| Importer 15 | 0.164737 | 0.143458 | 0.482578 | 0.611514 | 1.402287 | 1.198305393 | 90.0555215 |

**Supplementary Table 7:** Top fifteen (foreign) suppliers of antibiotics utilised in Tanzania between 2018 and 2021, with the proportion of each supplier in DID in four years and the cumulative contribution for each company.

| Suppliers | 2018 | 2019 | 2020 | 2021 | Total | Proportion of total DDI (%) | Cumulative % contribution |
| --- | --- | --- | --- | --- | --- | --- | --- |
| Supplier 1, China | 11.98331 | 3.45388 | 1.077391 | 0.292197 | 16.80677 | 14.36 | 14.36 |
| Supplier 2, India | 2.731248 | 1.581349 | 2.996334 | 1.57263 | 8.881561 | 7.59 | 21.95 |
| Supplier 3, Kenya | 0.193801 | 1.371094 | 0.696028 | 5.364539 | 7.625462 | 6.52 | 28.47 |
| Supplier 4, Kenya | 0.160403 | 3.281153 | 2.964335 | 3.578167 | 9.984058 | 8.53 | 37.00 |
| Supplier 5, Kenya | 0.915313 | 1.474415 | 2.85964 | 1.344799 | 6.594167 | 5.63 | 42.63 |
| Supplier 6, India | 0.185795 | 1.311286 | 1.165614 | 3.017761 | 5.680456 | 4.85 | 47.49 |
| Supplier 7, India | 0.571838 | 0.359887 | 2.322161 | 1.172949 | 4.426835 | 3.78 | 51.27 |
| Supplier 8, India | 0.358325 | 0.462936 | 1.478326 | 2.053054 | 4.352641 | 3.72 | 54.99 |
| Supplier 9, Kenya | 3.539278 | 0.218147 | 0.358759 | 0.063429 | 4.179613 | 3.57 | 58.56 |
| Supplier 10, India | 0.607576 | 0.62307 | 0.77138 | 1.735099 | 3.737125 | 3.19 | 61.76 |
| Supplier 11, India | 0.619548 | 0.425639 | 1.864228 |  | 2.909415 | 2.49 | 64.24 |
| Supplier 12, India | 0.760897 | 0.729129 | 0.559022 | 0.656169 | 2.705217 | 2.31 | 66.55 |
| Supplier 13, India |  | 0.83642 | 0.705891 | 1.047714 | 2.590025 | 2.21 | 68.77 |
| Supplier 14, India | 0.383732 | 0.805343 | 0.555065 | 0.608778 | 2.352918 | 2.01 | 70.78 |

**Supplementary Table 8:** Predicted Defined Dose per 1000 inhabitants per day (DID) from 2022 through 2027 from autoregressive integrated moving average ARIMA (0, 1, 0) model.

| Year | Actual<br>Daily<br>Defined<br>Dose per<br>1000<br>inhabitants<br>per day<br>(DID) | Predicted<br>DID | Lower<br>Control<br>Limit (LCL)<br>of DID | Upper<br>Control<br>Limit<br>(UCL) DID |
| --- | --- | --- | --- | --- |
| 2018 | 30.398308 |  |  |  |
| 2019 | 22.536254 | 31.305503 | -4.05338 | 66.66438 |
| 2020 | 30.968058 | 23.443449 | -11.9154 | 58.80233 |
| 2021 | 33.119892 | 31.875253 | -3.48363 | 67.23413 |
| 2022 |  | 34.027087 | -1.33179 | 69.38597 |
| 2023 |  | 34.934281 | -15.0707 | 84.93929 |
| 2024 |  | 35.841476 | -25.4019 | 97.08485 |
| 2025 |  | 36.748671 | -33.9691 | 107.4664 |
| 2026 |  | 37.655865 | -41.409 | 116.7207 |
| 2027 |  | 38.56306 | -48.0482 | 125.1743 |

## Notes

### Competing Interest Statement

The authors have declared no competing interest.

### Funding Statement

This study did not receive any funding.

